# Interaction of intestinal bacteria with human rotavirus during infection in children

**DOI:** 10.1101/2020.11.17.20233171

**Authors:** Roberto Gozalbo-Rovira, Antonio Rubio-del-Campo, Cristina Santiso-Bellón, Susana Vila-Vicent, Javier Buesa, Susana Delgado, Natalia Molinero, Abelardo Margolles, María Jesús Yebra, María Carmen Collado, Vicente Monedero, Jesús Rodríguez-Díaz

## Abstract

The gut microbiota has emerged as a key factor in the pathogenesis of intestinal viruses, including enteroviruses, noroviruses and rotaviruses (RV), where stimulatory and inhibitory effects on infectivity have been reported. With the aim of determining whether members of the microbiota interact with RV during infection, a combination of anti-RV antibody labelling, fluorescence-activated cell sorting and 16S rRNA amplicon sequencing was used to characterize the interaction between specific bacteria and RV in stool samples of children suffering diarrhea produced by G1P[8] RV. The genera *Ruminococcus* and *Oxalobacter* were identified as RV binders in stools, displaying enrichments between 4.8 to 5.4-fold compared to samples non-labelled with anti-RV antibodies. *In vitro* binding of the G1P[8] Wa human RV strain to two *Ruminococcus gauvreauii* human isolates was confirmed by fluorescence microscopy. Analysis in *R. gauvreauii* with antibodies directed to several histo-blood group antigens (HBGA) indicated that these bacteria express HBGA-like substances at their surfaces that can be the target for RV binding. Furthermore, *in vitro* infection of Wa strain in differentiated Caco-2 cells was significantly reduced by incubation with *R. gauvreauii*. These data, together with previous findings that had shown a negative correlation between *Ruminococcus* levels and antibody titers to RV in healthy individuals, suggest a pivotal interaction between this bacterial group and human RV. These results reveal likely mechanisms on how specific bacterial taxa of the intestinal microbiota could negatively affect RV infection and open new possibilities for anti-viral strategies.

## Introduction

Despite vaccination, group A rotavirus (RV) continues to be the leading etiologic agent of viral gastroenteritis in infants and young children worldwide [1] and is responsible for an estimated 130,000 deaths each year, mostly in developing countries [2].A traditional dual classification system of group A RV based on virion outer capsid proteins establishes more than 36 different G serotypes (depending on VP7 glycoprotein) and 51 P-types (depending on VP4 protease-sensitive protein) [3,4]. However, G1P[8], G2P[4], G3P[8] and G4P[8] RV are prevalent (over 90%) in most countries [5]. RV-host attachment is mediated by binding of the N-terminal portion of VP4 (VP8* fragment) to glycoconjugates at the cell surface. Diverse interactions between histo-blood group antigens (HBGA) and VP8* have been described and, recently, the reported differences in the recognition of the fucosylated secretory H type-1 antigen versus its non-fucosylated precursor in the predominant P[8] group explain the altered incidence of RV infections in secretor and non-secretor individuals [6,7].

In the complex gut ecosystem where RV develop, several interplays between the resident microbiota, the host glycobiology (e.g. mucosal HBGA) and enteric viruses have been described [6]. The classical view establishes that the gut microbiota protects against intestinal viral infections, but recent evidences also demonstrated a positive role for the microbiota in viral infection. Polioviruses and other intestinal viruses rely on the intestinal microbiota for infection by exploiting microbial-derived substances (e.g. lipopolysaccharide and peptidoglycan) to increase virion stability or to enhance attachment to host cells [8]. According to this, reduced viral infection is observed in gnotobiotic or antibiotic-treated animal models [8–11], a situation that has also been observed for RV in the mouse model [11]. Some probiotic and commensal gut bacteria have the ability to *in vitro* bind RV and human noroviruses [12–15], and HBGA-like substances have been detected in the surface of enteric species such as *Enterobacter cloacae* [13], *Enterobacter faecium, Klebsiella* spp., *Citrobacter* spp. and *Hafnia alvei* [16]. The bacterium *E. cloacae* enhances norovirus infectivity in gnotobiotic mice and in an *in vitro* model of infection in human B cells [10]. However, its application in gnotobiotic pigs reduced norovirus infectivity [17] for which some controversy still exists in this respect. The dual role of the microbiota in enteric virus infectivity (promoting or restricting) suggests that some microorganisms can be considered as risk factors while other can lead to protection against infection. In agreement with this concept, diverse bacterial groups have been correlated to diminished or increased antibody titers (reflecting previous infections) against RV and norovirus [18]. An important breakthrough has been recently achieved after identifying that segmented filamentous bacteria (SFB), a group of microorganisms present in rodents and other vertebrates, and intimately associated to the intestinal epithelium, protect mice against RV [19].

Due to the increasing evidence of gut microbiota implication in RV infection, the aim of this work was to investigate which bacteria interact with RV during natural infant infections and their likely role in the process.

## Results

### Determination of rotavirus binding bacteria by 16S rDNA sequencing

By using FACS coupled with FITC-labelled anti-RV antibody, we identified the bacteria interacting with RV in stool samples from five children suffering RV diarrhea clinically diagnosed as originated by the G1P[8] genotype. Total DNA was isolated from both sorted bacterial sub-populations (those RV-positive and -negative) and their microbial composition was determined by 16S rDNA sequencing. The information at the genus level was selected to analyze the differences in relative abundances of each bacterial group in the RV binding and non-binding bacteria. *Ruminococcus* genus was identified as a RV binder with percentages of abundance in the fluorescent versus the non-fluorescent bacterial groups showing a ratio > 5, followed by *Oxalobacter*, which presented a ratio > 4 (Fig. 1). However, variability between samples was very high and the differences were mainly due to individual samples. In particular, samples from two different individuals accounted for most of the differences in *Ruminococcus* and *Oxalobacter* in the detected microbiotas.

**Figure 1.**
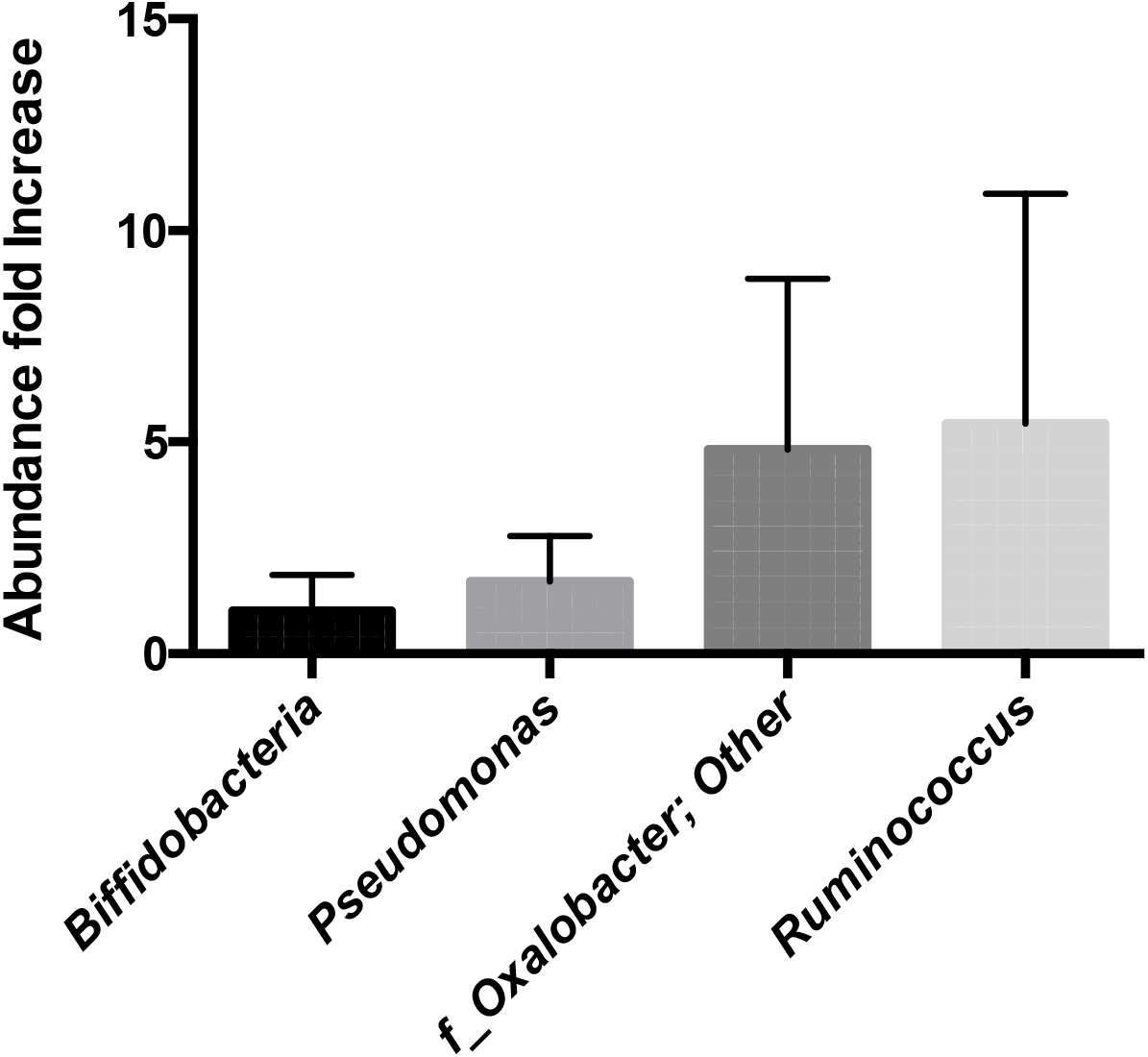
Increase in the relative abundance of several bacterial taxa in the fluorescent channel of FITC-labelled bacteria (rotavirus-bound) compared to non-labelled bacteria.

### *Ruminococcus gauvreauii* binds rotavirus *in vitro*

The fact that sequencing results revealed *Ruminococcus* as a bacterial genus with potential for RV binding attracted our attention owing to two different reasons: i) this genus was previously linked to lower anti-RV antibody titers in adult humans [18] and ii) the species *Ruminococcus gauvreauii* has been recently isolated from the human gall bladder [20]. We therefore analyzed the RV-*Ruminococcus* interaction using two strains of *R. gauvreauii*: the type-strain 19829, isolated from human feces, and the IPLA-NM1 strain, a human bile isolate [20]. As a RV strain we utilized Wa, a G1P[8] culture-adapted human RV with the same genotype than the pathogen present in the selected diarrheal samples. Both bacterial strains were able to bind Wa RV as determined by fluorescence microscopy. The binding of RV to *R. gauvreauii* DSM-19829 strain is shown in Fig. 2. Interestingly, adding RV to the bacterial samples also favored a certain degree of bacterial clumping, indicating that bacteria agglutination can be induced with RV (data not shown).

**Figure 2.**
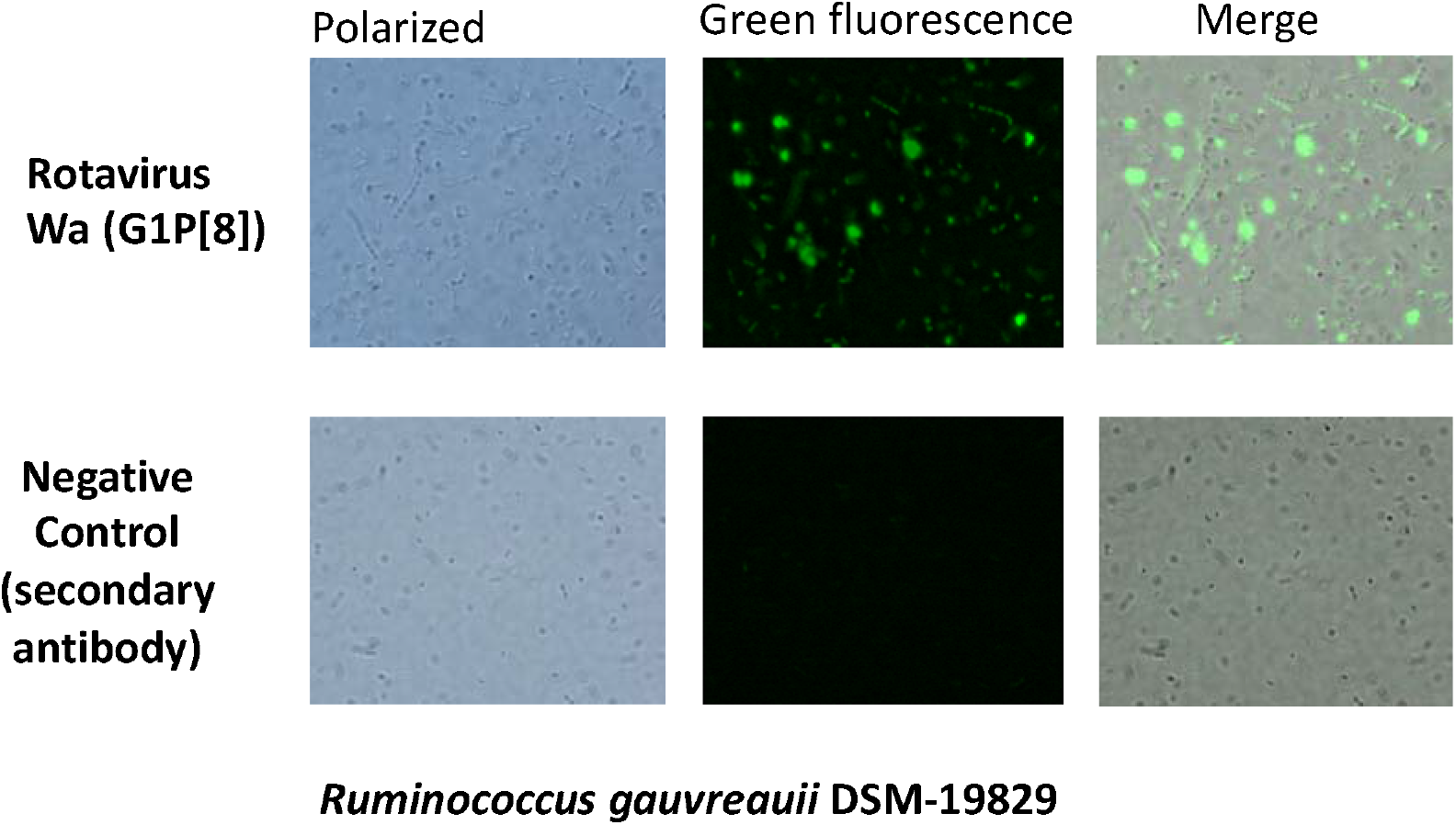
Microscopy images showing the interaction between bacteria from the genus *Ruminococcus* and RV by fluorescence microscopy. Bacteria were incubated with the G1P[8] rotavirus strain Wa or without rotavirus (control) and images were collected in the polarized field and in green florescence. Merged images from both fields are also presented.

### *R. gauvreauii* expresses HBGA-like substances on its surface

An ELISA assay was settled up to determine whether RV-binding *R. gauvreauii* expresses HBGA-like substances on its surface, providing a mechanism for bacteria-RV physical interaction (Fig. 3). Interestingly, the antibodies against the blood group A and B as well as against the H-antigen and the Lewis^a^ antigen resulted positive for both *R. gauvreauii* strains tested (DSM-19829 and IPLA-NM1), as well as for the positive control *E. cloacae* ATCC 13047. No signal was detected with the anti-Lewis^b^ antibody. The performance of this antibody was confirmed in an ELISA with saliva from a Le^b^ positive secretor individual as previously described [21], confirming that the bacteria did not express Le^b^-like substances on their surfaces. Interestingly, the ELISA results showed higher signals for some HBGA in *R. gauvreauii* strains compared to *E. cloacae*.

**Figure 3.**
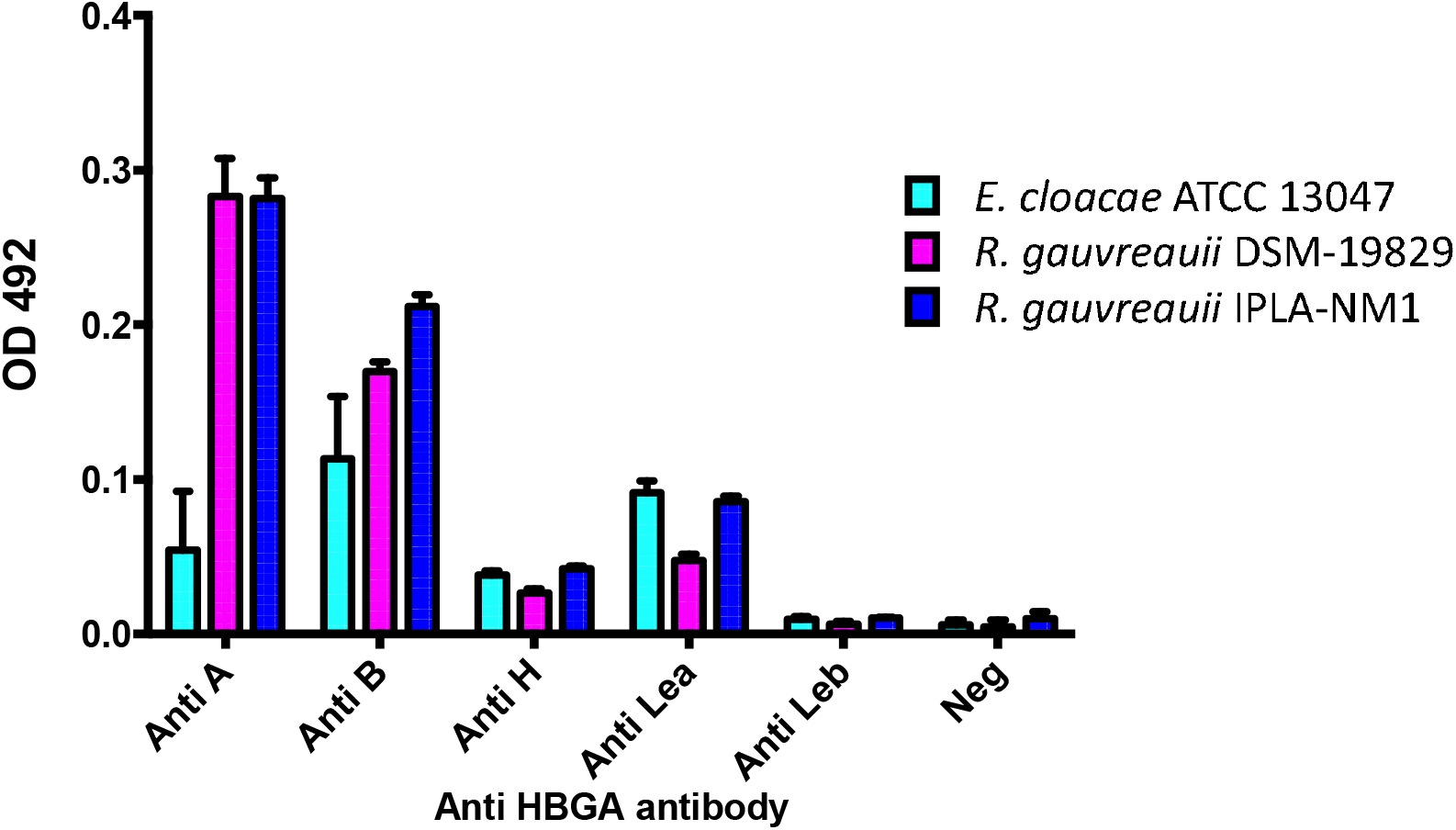
HBGA-like substances on the *Ruminococcus* surface detected by ELISA. *Ruminococcus gauvreauii* strains (DSM-19829 in purple and IPLA-NM1 in blue) and *Enterobacter cloacae* (positive control in light blue) were recognized by anti-A, anti-B, anti-H and anti-Lewis^a^ antibodies indicating that these bacteria express HBGA-like substances on their surfaces. Neg.: negative control (no antibody)

### *R. gauvreauii* interferes with rotavirus infection *in vitro*

Once the interaction between *R. gauvreauii* and RV was confirmed the antiviral properties of this bacterium were tested. Preincubation of Wa strain with *R. gauvreauii* followed by infection in Caco-2 cell did not produce a reduction in RV infectivity. An infection assay was then carried out in which the bacteria were pre-incubated with the cell monolayers before RV infection to simulate a natural infection, where the bacteria are pre-existing in the mucosal surface. Under these conditions it was determined that *R. gauvreauii* IPLA-NM1 attaches to the Caco-2 cells differentiated monolayer (Fig. 4A) and a 3-fold decrease in Wa RV infectivity (measured as viral genome equivalents/mL of cell culture supernatants after infection) was found, demonstrating an anti-RV activity for this bacterium (Fig. 4B).

**Figure 4.**
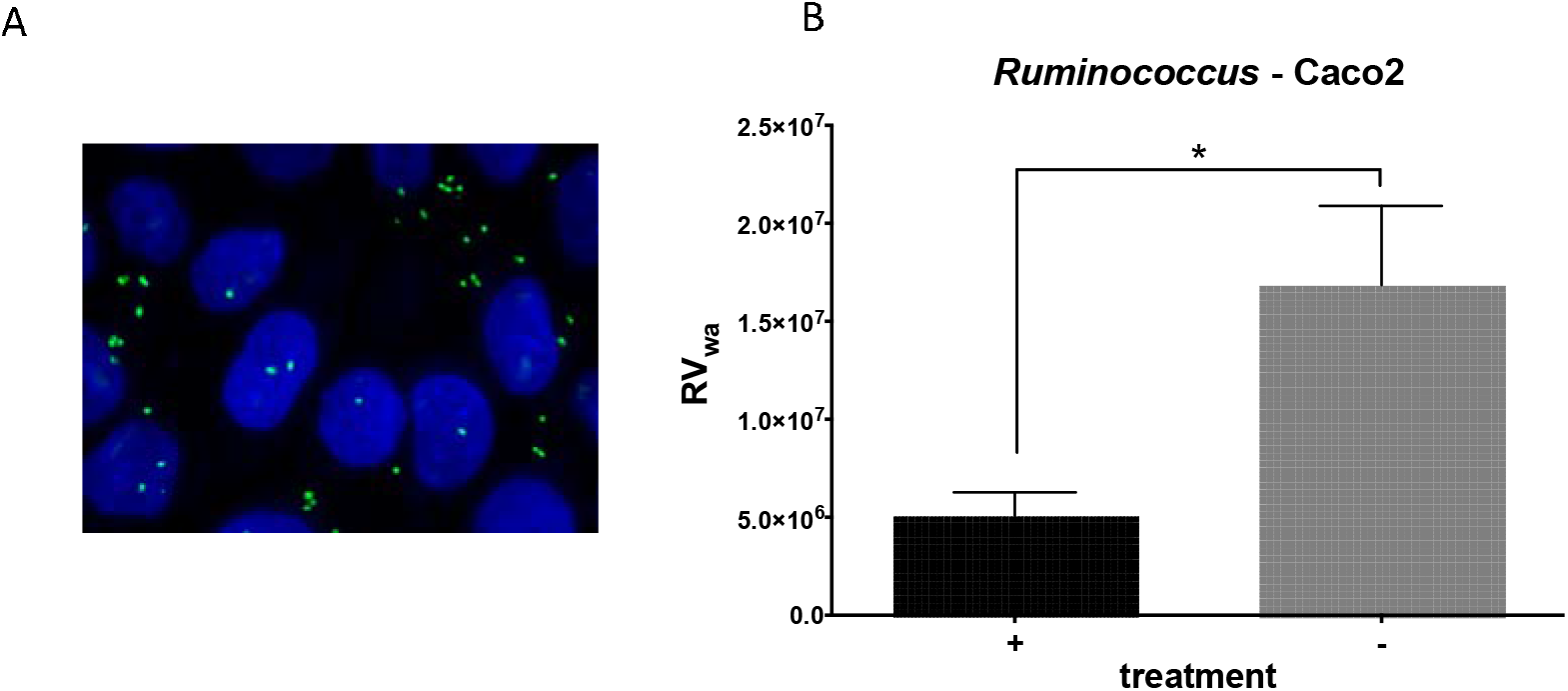
*Ruminococcus gauvreauii* and *in vitro* Wa infectivity. A) *R. gauvreauii* IPLA-NM1 cells bound to the Caco-2 monolayer. Cells and bacteria were stained with DAPI and several Z-stack images were taken and processed as described in material and methods. Caco-2 nuclei are in in blue, while *R. gauvreauii* cells are in green; B) Inhibitory effect of preincubation of *R. gauvreauii* on Wa rotavirus infection measured as reduction in viral genome equivalents in supernatants of Caco-2 cells after infection.

## Discussion

Since the demonstration that the microbiota plays a key role in the infection of virus targeting the intestine [8,22] many efforts have been performed to determine the role of gut bacteria in the relevant gastroenteritis-producing viruses, rotavirus and norovirus [6]. Unfortunately, despite the importance of this new concept, most of the evidences on the contribution of bacteria to RV infectivity have been achieved in cellular and animal models. Correlations between the abundance of several bacterial taxa and diminished antibody titers to norovirus and RV in adults have been reported [18], but data from infections in the relevant children group are still scarce and mainly derived from the results of seroconversion after vaccination with RV vaccine strains [23,24]. In this work we contribute to filling this gap through the utilization of clinical samples from children with acute RV diarrhea. Using these samples we have identified *Ruminococcus* and *Oxalobacter* as two bacterial genera interacting with RV during diarrhea in two separated samples. One objection that could be argued against the idea that the intestinal bacteria regulate the infectivity of RV is related to the different compartmentalization of RV replication and the intestinal microbiota. It is known that gut microbiota mainly resides in the colon, while RV replication occurs in the small intestine [3], where bacterial numbers and diversity is much lower compared to colon [25]. *Ruminococcus* and *Oxalobacter* are strictly anaerobic bacteria associated to the intestinal tract of mammals and they can be isolated from human feces, indicative of a colonic habitat [26,27]. *Ruminococcus* species have been associated to the important function of degrading resistant starch from food [28,29] and they are important in the maturation of the intestinal microbiota during human development [30], while O*xalobacter* is important for the metabolism of calcium oxalate at the intestine [31]. However, *Ruminococcus* has proved to reside also in the small intestine and gallbladder, as it can be isolated from bile [20]. The discovery of *Ruminococcus*-RV interaction also coincided with the fact that intestinal *Ruminococcus* levels were shown to negatively correlate with IgA titers against RV [18]. We demonstrated P[8] RV surface binding in *R. gauvreauii*. This surface attachment can be at the basis for the observed inhibitory effect of *R. gauvreauii* on the *in vitro* infectivity of Wa RV strain. It was previously shown that enteric bacteria such as *E. cloacae* expresses A-, B- and H-like antigens on its surface and is able to bind norovirus [13]. The same pattern of HBGA-like antigens was detected on the *R. gauvreauii* surface. The predominant P[8] RV interacts with H type-1 antigen and A-type HBGA, while Lewis antigens were reported not to fit the binding site for these antigens [32]. However, a recent report pointed to a second carbohydrate binding pocket for Lewis b in this viral genotype [33]. H type-1 antigen and A-type HBGA binding would provide a mechanism by which RV attach to the *R. gauvreauii* surface and suggest that virus-bacteria adhesion could produce a sequestration of viruses that impair viral infectivity. However, the fact that in our model of infection we only detected an inhibitory effect when *R. gauvreauii* was added to cultured cells prior to RV infection points to alternative mechanisms where bacterial priming of host cells is needed for reducing RV infection. A similar situation was described for the effect of *Lactobacillus* and *Bifidobacterium* in Wa [34,35] or bovine RV infection in culture cells [36]. However, a direct RV-bacteria interaction cannot be excluded. It has been recently shown that the chronic RV infection usually present in immunocompromised mice lacking B and T cell (Rag1-KO mice) can be prevented through ileal colonization by segmented filamentous bacteria (SFB; Candidatus Arthromitus) [19]. This protective effect does not involve immunoregulatory mechanisms triggered by SFB but probably depends on two non-mutually exclusive mechanisms: direct reduction of RV infectivity as a result of contact with SFB and enhanced enterocyte turnover. A recent study in the mouse model also evidenced a different role in RV infectivity for some bacterial groups. Infection by RV produces a shift in nutrient availability in the gut and induces goblet cells to release mucin, which promotes growth of mucin-degraders *Bacteroides* and *Akkermansia* members in mice ileum. Consumption of mucin by these bacteria may, in turn, decrease the protection exerted by mucin against RV infection [37]. It is not known at this stage whether high *Ruminococcus* levels would represent a protective factor against RV infection in children and, in fact, samples where *Ruminococcus* were detected derived from children suffering diarrhea. Nevertheless, *Ruminococcus* members are frequently found in the feces of children less than five years of age [38]. In a recent study, the microbiota in the feces from children suffering rotavirus diarrhea and from healthy children were compared, and *Ruminococcus* was found significatively more abundant in healthy children, reinforcing the idea of a possible antiviral role of this bacterial genus [39] confirming previous RV-microbiota correlation studies [18].

More studies are needed to unravel the contribution of specific microbiota taxons to RV infection in humans. Some works have pointed to the relevance of differences in intestinal microbiota composition on the lack of efficacy (low vaccine take) of RV vaccines, a live viral vaccine, in low-income and middle-income settings [23,24], but this issue is far from being solved and other possible contributing factors have been hypothesized [40]. Nevertheless, the bacteria-RV interacting pair here reported appears to be biologically relevant, since the interaction was detected in natural infections and a correlation *Ruminococcus*-RV susceptibility was previously evidenced in human samples [18]. Our data constitute new insights on the bacteria-enteric viruses’ interplay during infection and they could open new avenues for exploring innovative strategies to prevent and treat RV infections.

## Materials and methods

### Rotavirus detection and genotyping

Stool samples from children younger than 5 years with diarrhea attended in 2015 at the Hospital Clínico Universitario de Valencia that resulted positive for RV by immunochromatography (Certest Biotec) were selected for genotyping. The genotyping of G and P genotypes of samples was performed following the European Network EuroRotaNet procedures (www.eurorotanet.com). Five stool samples with the G1P[1] genotype were selected and stored at −80°C for further processing.

### Ethics statement

This study was conducted with the approval of the Ethics Committee of the Hospital (code F-CE-GEva-15; 26 March 2015), and informed written consent was obtained from patients’ parents/tutors before sample collection.

### Preparation of microbiota samples from stools and cell sorting

For detecting RV-binding bacteria we followed a strategy similar to that recently described to quantify the *in vitro* binding of noroviral virus-like particles to gut commensal bacteria [41]. Two-hundred mg of each stool sample were suspended in 1 mL of 0.9% saline solution (SS) by pipetting, followed by 10 seconds vortexing. The suspensions were centrifuged at 2,000 x g for 2 min to remove coarse materials and the supernatants were further centrifuged at 12,000 x g for 5 min to pellet the bacteria. The pelleted bacteria were washed twice in 1 mL SS. The washed pellets were resuspended in 300 *μ*L SS and fixed by adding 1,200 *μ*L of 4% paraformaldehyde overnight at 4ºC. After fixation, the cells were washed 3 times with SS and finally resuspended in 1 mL SS. One hundred *μ*L were reserved to study the sample autofluorescence in the flow cytometer and the other 900 *μ*L were further processed for RV immunostaining. The cells were pelleted and suspended in 150 *μ*L SS containing 5% bovine serum albumin (BSA) (Sigma-Aldrich) for 1 hour at 37 ºC to prevent nonspecific antibody binding. After blocking 1.5 *μ*L of FITC-labelled goat anti-RV antibody (ab31435, Abcam) was added and incubated for 1 hour at 37 ºC. After RV labelling, 1.5 *μ*L of 1 mg/mL solution of propidium iodide (Thermofisher) were added and incubated for 30 min at 37 ºC for nucleic acid staining. Finally, the cells were pelleted, washed twice with SS, and resuspended in 1 mL SS.

Bacterial cells were sorted using a cytometer FACSAria™III Cell and BD FACSDiva v6.1.6 analysis software (BD Biosciences, San Jose, CA) equipped with two-laser system at 488 nm and 633 nm (Supplementary Fig. 1). Cell sorting was made with a 70-μm nozzle. Microorganisms were determined using forward and side scatter parameters (FSC and SSC). FITC detector (530/30-nm-band-pass filter) was used to determine the levels of labelled bacteria (RV-binders) and propidium iodide, detector PE (585/42 nm-band-pass filter), was used to check for the presence of DNA. The parameter settings used were SSC signal at 200 units as the threshold to define an event to be counted, voltages of FSC 16, SSC 308, FITC 350, PE 455.

### Sorted fractions DNA extraction and 16S rDNA sequencing

Total DNA was isolated from the sorted bacteria by using the MasterPure Complete DNA & RNA Purification Kit (Epicentre) with a previous enzymatic step with 20 mg/mL lysozyme (Roche) and 10 U/mL mutanolysin (Sigma) at 37 ºC for 1 hour followed by glass bead beating (0,17 mm diameter) for 1 minute at 4ºC. The obtained DNA was amplified with the TruePrime WGA kit (Sygnis) following the instructions of the manufacturer. Total DNA concentration was measured and normalized using a Qubit® 2.0 Fluorometer (Life Technology, Carlsbad, CA, USA). Subsequently, the V3-V4 region of the bacterial 16S rDNA gene was amplified by PCR using Illumina adapter overhang nucleotide sequences following Illumina protocols. After 16S rDNA gene amplification, the mutiplexing step was performed with the Nextera XT Index kit (Illumina). Amplicons were checked with a Bioanalyzer DNA 1000 chip and libraries were sequenced using a 2×300pb paired-end run (MiSeq Reagent kit v3) on a MiSeq-Illumina platform (FISABIO Sequencing Service, Valencia, Spain). Controls during PCR amplification were also included and sequenced.

### Bioinformatics and statistical analysis

Raw reads were searched for residual adaptors using the program Trimmomatic [42]. Artifacts, quality check and quality trimming were performed with prinseq-lite program [43]. R1 and R2 fastq reads were then joined using overlapping reads with the program FLASH [44]. The quality filtered sequences were checked for chimera, and the non-chimeric sequences were processed using a QIIME pipeline (version 1.9.0) [45]. The sequences were clustered at 97% of identity into operational taxonomic units (OTUs), and representative sequences were taxonomically classified based on the Greengenes 16S rRNA gene database (version 13.8). Sequences that could not be classified to domain level, or classified as Cyanobacteria and Chloroplasts, were removed from the dataset. Subsequently, to study the RV-microbiota interaction patterns, the threshold used for including the genera was 0.5 % or greater in relative abundance in either the RV positive or negative fractions. The abundance proportions of a given genera were log-transformed before calculating the ratio between the RV positive or RV negative fractions, resulting in the RV index. The index (calculated according the formula log(RV++/RV-)) score reflects the degree of virus responsiveness and interaction to the specific microbial members, where the positive values represent the genera predominantly found to interact closely with RV while the negative values the bacterial genera predominantly not interacting with RV. Calypso software (http://cgenome.net/calypso/) was used with total sum normalization (TSS) for the statistical analysis.

The dataset supporting the conclusions of this article is available in the NCBI’s Sequence Read Archive (SRA) repository, BioProject ID PRJNA676006 (http://www.ncbi.nlm.nih.gov/bioproject/676006).

### Bacterial strains and culture conditions

*R. gauvreauii* DSM-19829 and *R. gauvreauii* IPLA-NM1 [20] strains were used in this study. *R. gauvreauii* DSM-19829 was purchased from the DSMZ Collection, and IPLA-NM1 strain was isolated from a human bile sample [20]. They were grown in Gifu Anaerobic Medium (Nissui) supplemented with 0.25% (w/v) L-cysteine (Sigma-Aldrich) (named GAMc) incubated at 37°C for 72 h in a Whitley MG500 anaerobic cabinet (Don Whitley Scientific) under a 10% H2, 10% CO2, and 80% N2 gas atmosphere. The two strains were routinely maintained by growing in 2% agar GAMc plates under anaerobic conditions.

### Production of rotavirus infectious viral particles

The RV Wa strain, G1P[8] was cultured in MA104 cells and purified as previously described (Gozalbo-Rovira et al., 2019). Briefly, 10 confluent 150-cm^2^ flasks (approximately 1.5 ×107 cells/flask) were infected with Wa strain at a multiplicity of infection (MOI) ≤ 0.1. One hundred mL of medium with 1,5×10^8^ virus/mL were obtained and the viral particles were concentrated by pelleting at 160,000 × g for 1 h at 4°C in a SW 41 rotor (Beckman). The viral pellet was resuspended in TNC buffer (20 mM Tris-HCl pH 8.0, 100 mM NaCl, 1 mM CaCl2) for obtaining triple-layered particles and the particles were visualized by transmission electron microscopy after negative staining with phosphotungstic acid.

### *Ruminococcus*-rotavirus binding assay

Cells from *Ruminococcus gauvreauii* DSM-19829 and *R. gauvreauii* IPLA-NM1, an isolate from human small intestine [20] were washed and diluted in PBS to an OD595 of 1. The cells were incubated with RV Wa at a concentration of 100 *μ*g/mL of total viral antigen for 1 hour at 37ºC with gentle agitation to allow the bacteria interact with the virus in a final volume of 200 *μ*L. After this period the cells were centrifuged and washed two times with PBS to remove unbound RV. The cells were resuspended in 200 *μ*L of fixing solution (4% formaldehyde in PBS) and incubated for 15 min at 37 ºC. After fixing, the bacterial suspensions were spread on glass microscope slides and air dried. The slides were blocked for 1 h at room temperature in PBS containing 1% BSA and the viruses were detected by incubation with mouse anti-RV IgG antibody raised in the laboratory, followed by AlexaFluor488-labelled goat anti-mouse IgG antibody (1:400, Molecular Probes). The preparations were mounted with 10 *μ*L of ProLong Gold Antifade Reagent (Life Technologies) and visualized with an Eclipse 90i fluorescence microscope (Nikon). Controls without the virus and without the primary antibody (mouse anti-RV) were included.

### HBGA ELISA on bacterial cells

An ELISA assay was settled up to determine whether the *Ruminococcus* strains expressed HBGA-like substances in their surface. ELISA plates (Costar) were coated with the two *R. gauvreauii* strains (DSM-19829 and IPLA-NM1). *Enterobacter cloacae* ATCC 13047 (grown in nutrient broth at 37 ºC overnight with shaking) was used as positive control since it was previously determined that it expresses HBGA-like substances at its bacterial envelope [13]. The bacterial cells were washed three times in PBS and diluted to an OD595 of 1 in PBS. The wells of an ELISA plate were coated with 100 *μ*L of the bacterial suspensions by incubation at 4ºC overnight. After coating, the wells were washed three times with 200 *μ*L PBS containing 0.05 % Tween 20 (PBS-T) and blocked with PBS-T containing 3% BSA for 1 hour at 37ºC. After blocking, the plate was incubated for 1 h at 37ºC with monoclonal antibodies directed to HBGA (anti-A and anti-B, Diagast); anti-H, anti-Le^a^ and anti-Le^b^, Covance) diluted 1:100 in PBS with 1% BSA. After three washes, horseradish peroxidase goat anti-mouse IgG (Sigma) diluted 1:2,000 in PBS-T plus 1% BSA was added and incubated for 1 hour at 37ºC. After three washes with PBS-T the reactions were developed with o-phenylenediamine dihydrochloride (OPD-Fast) (Sigma), stopped with 2M H_2_SO_4_, and recorded at 492 nm. The cut-off value was defined as a threefold increase in absorbance value compared to the negative control (cells incubated only with the secondary antibody). All the experiments were performed in triplicate.

### Rotavirus infection blocking assay with *R. gauvreauii*

Caco-2 cells were placed in 96 wells cell culture plates (Costar) at 5×10^4^ cells/well in MEM medium containing 10% foetal bovine serum (FBS), 1% non-essential amino acids and 1 X penicillin/streptomycin and allowed to differentiate for 11 days in a CO_2_ incubator at 37ºC with culture medium changes every three days. After differentiation, cell monolayers were washed with serum-free medium and two different infection strategies were applied: i) Wa RV (1.5×10^4^ genome equivalents that were previously activated by incubation in serum-free medium with type IX trypsin (Sigma) at 10 *μ*g/mL for 30 min at 37 ºC) were mixed with *R. gauvreauii* IPLA-NM1 at an OD595 of 0.1 in MEM medium and incubated for 60 min prior addition to the Caco-2 cells; ii) *R. gauvreauii* IPLA-NM1 cells were added at OD_595_ of 0.01 to Caco-2 cells and incubated for 1 hour at 37ºC. Control wells without added bacteria were included. Caco-2 monolayers were washed three times with serum-free medium containing 1 *μ*g/mL type IX trypsin (Sigma) and infected with activated rotavirus (1.5×10^4^ genome equivalents / well). In both infection strategies, RV were kept in contact with the Caco-2 monolayers for 1 h at 37ºC. After this incubation period the viral inoculum was removed and the wells washed twice with serum-free medium and incubated with serum-free medium containing 1 *μ*g/mL type IX trypsin (Sigma) for 16 hours. After this period the cell monolayers were disrupted with PBS containing 1% Triton X-100 and RNA was extracted using the Nucleospin-RNA virus Kit (Macherey-Nagel) following the supplier instructions. The RV titers obtained after infection were calculated by RT-qPCR as previously described [46]. Bacterial adhesion to the Caco-2 monolayers was also visualized by fluorescence. After incubation of the bacterial suspension with the Caco-2 monolayer, the cells were fixed and stained with DAPI. Nine 1 *μ*m Z-stack images were taken at 100 x using an Eclipse 90i microscope (Nikon corporation, Japan). These image stacks were processed with Fiji (ImageJ 1.49q Software, National Institutes of Health, USA) and subdivided in two stacks, which contained basal visual information of the Caco-2 monolayer (mainly nuclei) and the bacteria located on top, respectively. Both sub-stacks were processed separately and finally merged in single images.

## Data Availability

The dataset supporting the conclusions of this article is available in the NCBI's Sequence Read Archive (SRA) repository, BioProject ID PRJNA676006

http://www.ncbi.nlm.nih.gov/bioproject/676006

## Figure legends

**Supplementary Figure 1.**
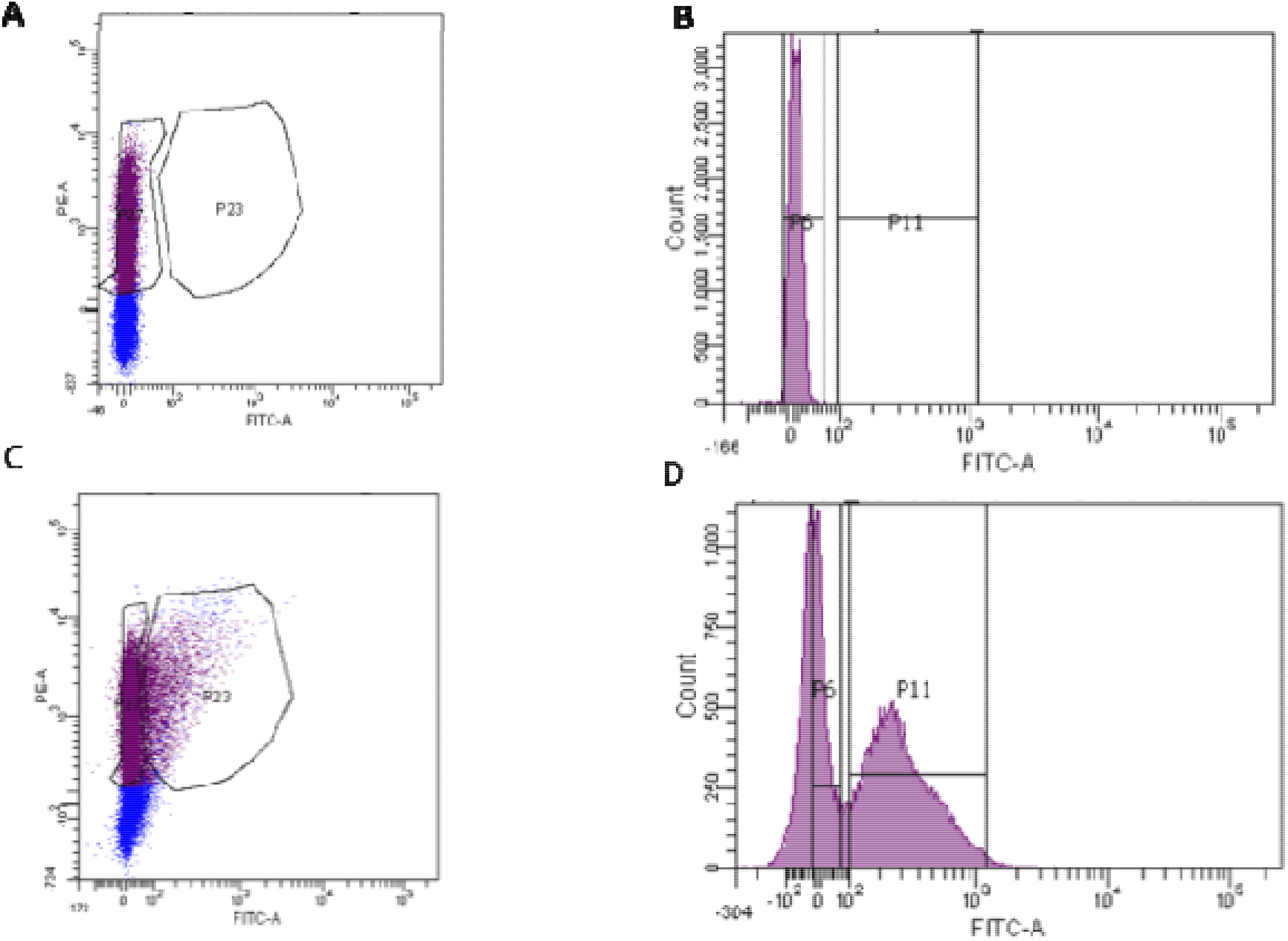
Fluorescence-activated cell sorting of rotavirus-bound fecal microbiota. Non-labelled bacteria from the stools of patients were subject to FACS for determining auto-fluorescence and establish the non-binding population (panels A and B). The same bacteria were labelled with FITC-anti-rotavirus and subject to FACS (panels C and D). The P6 population contained the non-FITC fluorescent bacteria and the P11 population contained the FITC fluorescent bacteria with bound rotavirus. The bacteria (labelled and non-labelled) were collected separately and analyzed by 16S rDNA sequencing.

## Acknowledgements

This work was supported by Spanish Government (Ministerio de Economía y Competitividad) grants AGL2017-84165-C2-1-R to MJY, AGL2017-84165-C2-2-R and RYC-2013-12442 to JRD. This work was also supported by Valencian Government grant IDIFEDER/2018/056. RGR is the recipient of a postdoctoral grant from the Valencian Government APOST/2017/037. CSB is the recipient of a predoctoral grant FPI from the Spanish Government RE2018-083315. SVV is the recipient of a predoctoral grant from the Valencian Government ACIF/2016/437. We thank J.M. Coll-Marqués for digital treatment of microscope images.

